# Characterizing Genetic Pathways Unique to Autism Spectrum Disorder at Multiple Levels of Biological Analysis

**DOI:** 10.1101/2024.06.07.24308616

**Authors:** Lukas S. Schaffer, Sophie Breunig, Jeremy M. Lawrence, Isabelle F. Foote, Andrew D. Grotzinger

## Abstract

Autism spectrum disorder (ASD) is a neurodevelopmental condition characterized by atypical patterns of social functioning and repetitive/restricted behaviors. ASD commonly co-occurs with ADHD and, despite their clinical distinctiveness, the two share considerable genetic overlap. Given their shared genetic liability, it is unclear which genetic pathways confer unique risk for ASD independent of ADHD. We applied Genomic Structural Equation Modeling (SEM) to GWAS summary statistics for ASD and ADHD, decomposing the genetic signal for ASD into that which is unique to ASD (*uASD)* and that which is shared with ADHD. We computed genetic correlations between *uASD* and 75 external traits to estimate genetic overlap between *uASD* and other clinically relevant phenotypes. We went on to apply Stratified Genomic SEM to identify classes of genes enriched for *uASD*. Finally, we implemented Transcriptome-Wide SEM (T-SEM) to explore patterns of gene-expression associated with *uASD*. We observed positive genetic correlations between *uASD* and several external traits, most notably those relating to cognitive/educational outcomes and internalizing psychiatric traits. Stratified Genomic SEM showed that heritability for *uASD* was significantly enriched in genes involved in evolutionarily conserved processes, as well as for a histone mark in the germinal matrix. T-SEM revealed 83 unique genes with expression associated with *uASD,* many of which were novel. These findings delineate the unique biological underpinnings of ASD which exist independent of ADHD and demonstrate the utility of Genomic SEM and its extensions for disambiguating shared and unique risk pathways for genetically overlapping traits.

## Introduction

Autism spectrum disorder (ASD) is a heterogenous neurodevelopmental disorder which occurs in >1% of the population [1, 2]. While the phenotypic presentation of ASD is highly variable, it is characterized by two core symptom domains: (i) impairments in social communication and interaction as well as (ii) repetitive, restricted patterns of behavior or interests [3]. Although the etiology of ASD involves an array of risk factors, extant literature has demonstrated a strong genetic component with heritability estimates from twin and family studies ranging between 64 and 91% [4–6]. A substantial proportion of this heritability (∼12%) is attributable to common genetic variation as evidenced by recent genome-wide association studies (GWASs) of ASD [2, 7].

ASD often presents alongside other neuropsychiatric conditions; the most frequent comorbidity is attention-deficit/hyperactivity disorder (ADHD) [8], a childhood-onset disorder characterized by symptoms of either inattention, hyperactivity, or both [3]. Indeed, conservative estimates suggest that one in every three children with ASD will also meet diagnostic criteria for ADHD [9]. Converging evidence indicates that a common genetic liability partially underlies risk for both disorders [10]. For example, observations from family studies find that ASD and ADHD tend to co-aggregate in families [11], and this co-aggregation is due, in part, to shared additive genetic influences [12, 13]. These findings are corroborated by molecular and statistical genetic studies, which have estimated moderate genetic correlations between ASD and ADHD [2, 10, 14], indicating a shared genetic architecture.

These findings illustrate a broad challenge of parsing disorder-specific biological pathways when two phenotypes are genetically and phenotypically correlated. These difficulties necessitate the need for multivariate genomic analyses capable of isolating the genetic variance that is unique to a specific trait. Here, we approach these challenges by leveraging Genomic Structural Equation Modeling (SEM) and its extensions to separate out the genetic signal unique to ASD from that which is shared with ADHD [15–17]. We model GWAS summary statistics for ASD and childhood-diagnosed ADHD using a Cholesky decomposition to derive a unique ASD (henceforth, *uASD*) latent factor, reflecting the residual genetic signal for ASD after removing genetic overlap with ADHD. We then apply downstream analyses to interrogate the genetic architecture of *uASD* at the genome-wide, functional, and gene-expression levels of analysis. Collectively, these analyses delineate the biological mechanisms that contribute specifically to the etiology of ASD and its associated symptoms, as opposed to those that may confer a broader spectrum of shared neurodevelopmental risk.

## Method

### Summary Statistics

Summary statistics for ASD were used from the most recent GWAS meta-analysis [2]. Briefly, the original GWAS included 13,076 cases and 22,664 controls from the Danish population-based cohort iPSYCH and 5,305 cases and 5,305 pseudo-controls (i.e., non-transmitted parental alleles) from family-based trio samples from the Psychiatric Genomics Consortium (PGC). Together, the meta-analysis totaled 18 381 ASD cases and 27 969 controls/pseudo-controls. GWAS summary statistics for childhood-diagnosed ADHD were utilized from the GWAS conducted by Rajagopal et al. (2022), which stratified ADHD cases by age of diagnosis. Here, we specifically utilize summary statistics for ADHD diagnosed in childhood due to its higher genetic overlap with ASD compared to persistent or adulthood-diagnosed ADHD [14, 18]. The childhood-diagnosed GWAS included 14 878 cases and 38 303 controls from the iPSYCH cohort.

To evaluate genetic overlap with other relevant phenotypes, we leveraged publicly available European-ancestry summary statistics for 77 external traits spanning domains of cognition, psychopathology, health/lifestyle behaviors, interpersonal relations, and physical activity. We used a SNP-based *h^2^ z*-statistic cutoff of 4, as recommended by the original linkage disequilibrium score regression (LDSC) developers [19], to limit our pool of external traits to those with interpretable genetic covariance. Based on this cutoff, 75 of our original 77 traits were carried forward for analysis. A comprehensive list of included external traits and relevant characteristics is reported in **Supplementary Table 1**.

### Genomic Structural Equation Modeling

Prior to analysis, all GWAS summary statistics underwent an identical set of QC filters using the *munge* function in the *GenomicSEM* R package. These filters included restricting analyses to HapMap3 SNPs and removing SNPs with a minor allele frequency (MAF) < 1% and imputation score (INFO) < .9 (when available). Once processed, these GWAS summary statistics were used as input for multivariable LDSC using the *ldsc* function within *GenomicSEM*, which produces the genetic covariance and sampling covariance matrices across included traits. The genetic covariance matrix includes the SNP-based heritability (*h^2^*) on the diagonal and the genetic covariances on the off-diagonal. The sampling covariance matrix contains squared standard errors (sampling variances) on the diagonal and the sampling covariances (sampling dependencies) on the off-diagonal that will arise in the presence of participant sample overlap. The sampling covariance matrix is estimated directly from the data using a block jackknife resampling procedure and allows for GWAS with varying degrees of power and sample overlap to be included in the same statistical model.

For binary traits, estimates were converted to the liability scale using the population prevalence and the sum of effective sample size across contributing cohorts [20]. For ASD, the effective sample size was estimated directly from the data. This is because the ASD GWAS used pseudocontrol subjects, which reduces power to detect GWAS associations, such that using the observed sum of effective sample size would produce downwardly biased estimates of heritability [21]. LDSC requires that estimates are produced within a single ancestry group as the LD weights used to estimate the regression model will vary across ancestral populations. Due to limited availability of data in other ancestral groups, GWAS statistics were limited to participants of European ancestry, and the LDSC model was estimated using the 1000 Genomes Phase 3 European LD scores. These scores excluded the major histocompatibility complex (MHC) due to complex LD structures in this region that can bias estimates.

The output from LDSC was used as input to all subsequent analyses in Genomic SEM. We began by fitting a Cholesky decomposition model to our observed variables based on GWAS summary statistics for ASD and childhood-diagnosed ADHD (henceforth, simply referred to as ADHD). Both ASD and ADHD were regressed onto a latent factor, *cADHD*, which represents the genetic variance of ADHD as well as the proportion of genetic variance for ASD that is shared with ADHD. ASD was additionally regressed onto *uASD*, representing the residual genetic variance that is unique to ASD after accounting for that which is shared with ADHD. By construction, *uASD* and *cADHD* were orthogonal (*r_g_* = 0). The genetic residual variances of ASD and ADHD were fixed to 0 so that all variance in the disorders was explained by the latent factors. At the genome-wide level, this model was expanded to compute the genetic correlations between *uASD* and each of our pre-selected 75 external traits (see **Supplementary Table 2**). In interpreting the statistical significance of each genetic correlation, we apply a strict Bonferroni-adjusted significance threshold (*p* < 6.7E-4).

### Stratified Genomic SEM

We applied Stratified Genomic SEM to identify enrichment for functional annotations (i.e., categories of genes) for *uASD*. We first ran multivariable Stratified LDSC (S-LDSC) to obtain genetic covariance estimates within each annotation. We originally included a total of 168 annotations for analysis. However, 12 of these annotations were excluded due to the model failing to converge (*n* = 10) or negative heritability estimates (*n* = 2). The remaining 156 were examined to ensure that none required a smoothing of the covariance matrix resulting in a *z*-statistic difference > 1.96 (as recommended by Grotzinger et al., 2022) before moving forward. Our final analysis included 49 annotations from the 1000 Genomes Baseline LD Version 2.2 [22], as well as neuronal and brain tissue annotations from DEPICT [23], gnomAD [24], GTEx v8 [25], and the Roadmap Epigenomics Project [26] (see **Supplementary Table 3** for full list of annotations). Using the *enrich* function within *GenomicSEM*, we then estimated enrichment for *uASD* within each annotation. Enrichment is calculated as the ratio-of-ratios. For the current analyses, the numerator of this ratio reflects the proportion of *uASD* genetic variance explained by an annotation (i.e., the within-annotation genetic variance for *uASD* divided by the total *uASD* genetic variance). The denominator of the ratio reflects the proportional size of the annotation (i.e., the number of SNPs in the annotation divided by the total number of SNPs analyzed across all annotations). The null for this enrichment ratio-of-ratios is 1, where values above 1 index functional annotations that account for a greater proportion of genetic variance in *uASD* than would be expected based solely on the proportional size of that annotation. Given the non-independent nature of functional annotations, we applied the FDR correction for multiple comparisons to the accompanying *p*-values using the *p.adjust* R package.

### Transcriptome-Wide SEM (T-SEM)

Transcriptome-wide SEM (T-SEM) was applied to identify patterns of gene expression associated with *uASD*. First, FUSION [27] was used to perform univariate transcriptome-wide association studies (TWASs) on both ASD and ADHD. We utilized functional weights for 13 brain tissue types from the Genotype Tissue Expression Project (GTEx v8, [28]), two dorsolateral prefrontal cortex weights from the CommonMind Consortium (CMC, [29]), and one set of weights for the prefrontal cortex from PsychEncode [30]. This resulted in 16 total functional weights from which we derived 73 412 genes with imputed expression data across different brain regions and tissues. These univariate FUSION outputs were then input into the *read_fusion* function in Genomic SEM.

The gene expression estimates were then added to the LDSC covariance matrix for ASD and ADHD, and the *userGWAS* function was used to estimate the effect of gene expression on both *uASD* and *cADHD*. Given the scope of the analyses, we specifically focus on the relationship between gene expression and *uASD*. Finally, to identify gene sets with significant enrichment in *uASD*, we used the *WebGestalt* package to conduct an overrepresentation analysis (ORA, [31]) on significant T-SEM hits. An FDR correction was used in interpreting significance of T-SEM and ORA results. We also carried forth a drug repurposing analysis on these hits using methods outlined by Grotzinger et al. 2023. As these analyses did not produce any findings, no results are reported below.

## Results

### Genome-Wide Results Reveal Genetic Correlates of uASD

ASD and ADHD were moderately genetically correlated (*r_g_* = .45, *SE* = .05). The Cholesky decomposition revealed that *cADHD* explained ∼20% of the genetic variance in ASD and *uASD* explained the remaining (∼80%) ASD genetic variance (see **Figure 1** for visual representation of the structural equation model and partitioning of genetic variance). The remaining analyses sought to clarify what genetically differentiates ASD from ADHD by examining associations with the *uASD* factor at multiple levels of analysis. We began by genetically correlating *uASD* with 75 external traits in order to evaluate the extent of genetic overlap with other phenotypes relevant to mental and physical health. By virtue of our statistical definition of *uASD,* these correlations represented associations extending above and beyond those with *cADHD*. A full list of external traits and relevant outputs is available in **Supplementary Table 2**. Our analyses revealed that *uASD* was significantly correlated with 20 external traits. These 20 correlations tended to span four primary phenotypic dimensions – cognition, psychopathology, physical movement, and interpersonal relations – that we review below (and in **Figure 2**, panels **A, B, C,** and **D**).

**Figure 1.**
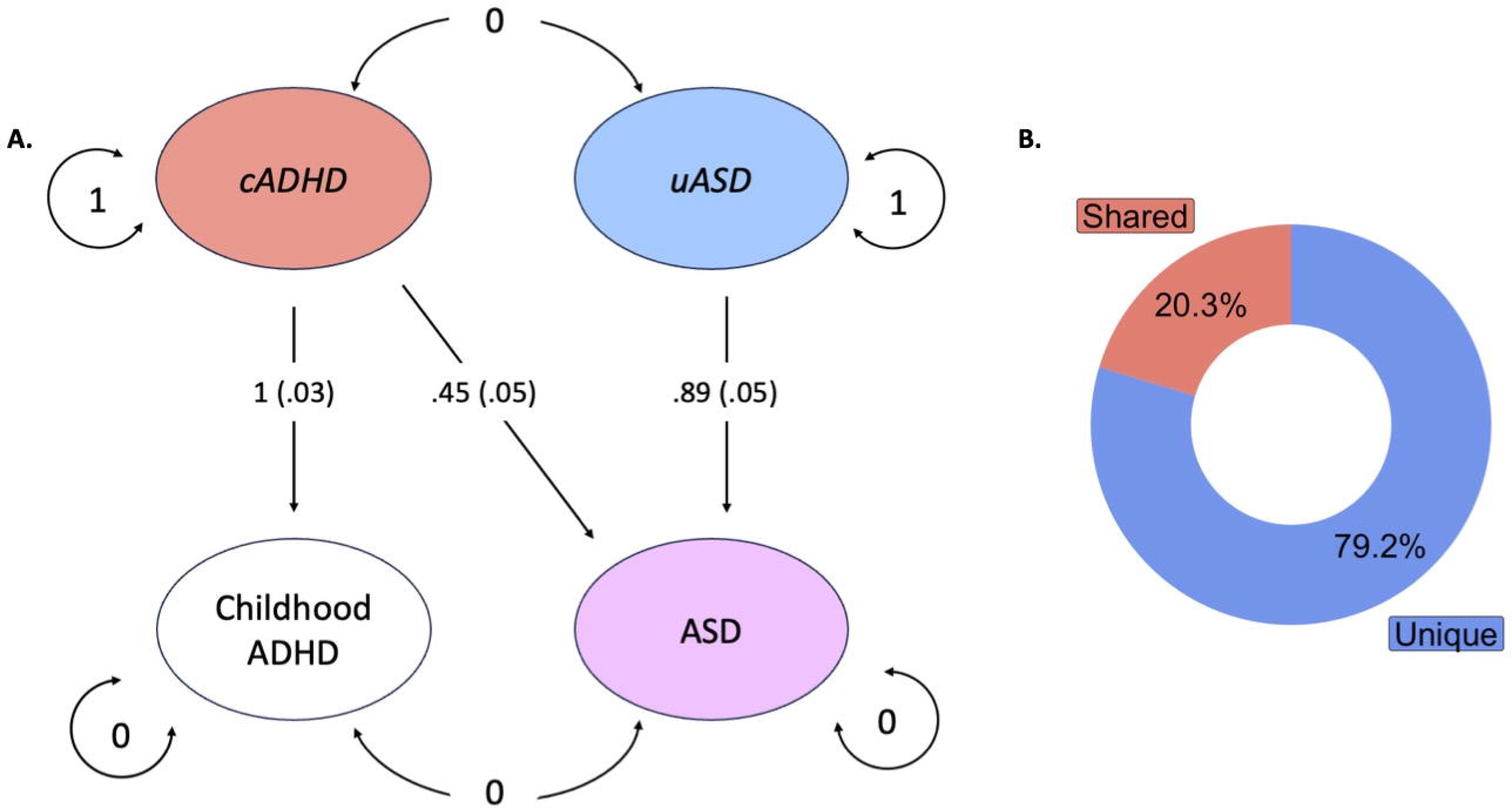
Genomic structural equation modeling to decompose ASD genetic variance. **(A)** Cholesky decomposition model producing *uASD*, a latent variable encompassing the genetic variance unique to ASD independent of ADHD, and *cADHD,* which captures the residual genetic variance of ASD (i.e., variance shared between ASD and ADHD) and the genetic variance of ADHD. **(B)** Donut plot showing the proportion of residual genetic variance unique to ASD (blue) and shared with childhood ADHD (red).

**Figure 2.**
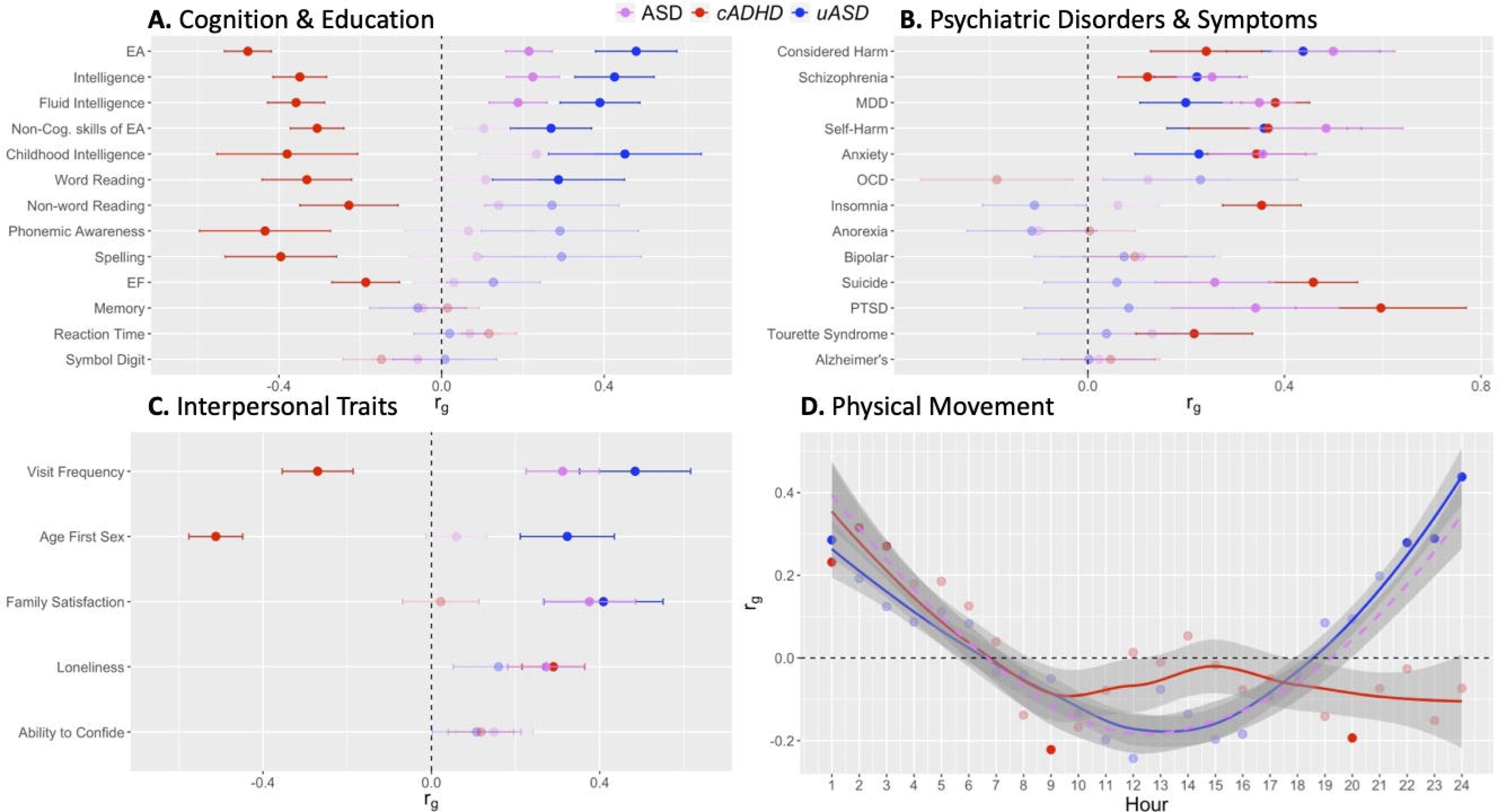
Genetic correlations between *uASD* and external traits. Genetic correlations between *uASD* (blue) and external traits for domains of cognition and education **(A)**, psychiatric disorders and symptoms **(B)**, and interpersonal traits **(C)**. Traits are sorted top to bottom by ascending *p-*value for the *uASD* correlation. **(D)** Genetic correlations between accelerometer-based average total hourly movement within the 24-hour day beginning at midnight (i.e., hour 1) and *uASD* and *cADHD*. Correlations are also shown between external traits and ASD (pink) as well as *cADHD* (red). Error bars represent 95% confidence intervals. Translucent points and error bars represent genetic correlations that did not surpass the Bonferroni-adjusted significance threshold. Panel D depicts a LOESS regression line used to visualize overall trends across individual point estimates. Shaded region around the regression line represents a 95% confidence interval. Dashed pink line represents the LOESS regression line for genetic correlations with ASD. EA = educational attainment, EF = executive function, MDD = major depressive disorder, OCD = obsessive-compulsive disorder, PTSD = post-traumatic stress disorder.

#### Cognition

Cognitive-related phenotypes demonstrated the most robust genetic correlations with the latent factor *uASD*, both in statistical significance and magnitude. We observed positive correlations between *uASD* and educational attainment (*r_g_*=.48, SE = .05), childhood intelligence (*r_g_* = .45, SE = .1), general intelligence (*r_g_* = .42, SE = .05), noncognitive skills of educational attainment (*r_g_* = .27, SE = .05), word reading (*r_g_* = .29, SE = .08), and verbal numerical reasoning (*r_g_* = .39, SE = .05). Interestingly, a number of these traits (childhood intelligence, noncognitive skills of educational attainment, and word reading) were not found to be significantly associated with ASD, indicating that *uASD* may be capturing additional genetic variance uniquely related to cognitive- and education-related traits. While we observed consistent associations with traits indexing intellectual abilities, we did not observe a statistically significant correlation between *uASD* and the trail-making task B, a well-established measure of executive functioning [32, 33]. Collectively these cognitive results indicate that the genetic component unique to ASD is specifically associated with more general cognitive processes independent of self-regulatory processes.

#### Psychopathology

Several psychiatric phenotypes, especially those falling within the internalizing spectrum, were positively and significantly associated with *uASD*. These included anxiety (*r_g_* = .22, SE = .07), major depressive disorder (MDD; *r_g_* = .20, SE = .05), self-harm (*r_g_* = .36, SE = .10), consideration of self-harm (*r_g_* = .44, SE = .08), and sensitivity to environmental stress and adversity (*r_g_*= .16, SE = .05). A positive genetic correlation was also observed between *uASD* and schizophrenia (*r_g_*= .22, SE = .04). These genetic correlations with psychopathology were similar in directionality and magnitude to those observed for ASD and *cADHD*. This suggests that, despite their high level of genetic overlap, ASD and ADHD each have unique genetic pathways that link them to other forms of psychopathology.

Given the shared signal between *uASD* and other psychiatric disorders, we went on to implement a follow-up model to examine the magnitude of unique genetic signal in ASD after partialling out genetic overlap with multiple psychiatric disorders. This involved running a multiple regression model with ADHD, MDD, anxiety, and schizophrenia as correlated predictors of ASD. The multiple regression model path diagram is visualized in **Supplementary Figure 1** and its output is provided in **Supplementary Table 3.** This model revealed that 74% of the residual genetic variance in ASD was unexplained by the other psychiatric disorders. This demonstrates that ASD is not merely a genetic amalgamation of ADHD and other psychiatric disorders, but has a sizable proportion of unique genetic variance distinguishing it from other forms of psychopathology.

#### Physical movement

We examined the genetic correlations with accelerometer data (i.e., physical movement) in 1-hour increments across a 24-hour period. This accelerometer data can be considered a useful indicator of atypical patterns of movement that may reflect disturbances in sleep and circadian rhythm, wherein these processes have been discussed as transdiagnostic risk factors for psychiatric and neurodevelopmental disorders [16, 34, 35]. We observed positive associations between *uASD* and movement at hours 0-1 (i.e., 12:00AM-1:00AM; *r_g_* = .29, SE = .08), 21-22 (i.e., 9:00PM-10:00PM; *r_g_* = .28, SE = .07), 22-23 (*r_g_* = .29, SE = .08), and 23-24 (*r_g_* = .44, SE = .08), indicating that this genetic overlap was restricted to movement during periods of early morning and late night. We also observed overlap between *uASD* and physical inactivity (*r_g_* for hours of moderate exercise = −.25, SE = .07).

#### Interpersonal relations

A select few traits relating to increased social behavior displayed genetic correlations with *uASD*. Notably, these positively associated traits tended to revolve around social relations with family members, such as family satisfaction (*r_g_* = .41, SE = .07) and the frequency of friend and family visits (*r_g_* = .48, SE = .07). Age of first sexual encounter was also positively genetically correlated with *uASD* (*r_g_* = .32, SE = .06), and this association was not observed with ASD.

### Functional Results Identify uASD Enrichment

Stratified Genomic SEM revealed four significantly enriched functional annotations for *uASD*. Three of these annotations reflected classes of genes implicated in evolutionarily conserved processes, including genes conserved in primates (*p* = 1.00E-7), and two annotations indexing genes conserved in mammals (*p* = 5.68E-4 and 9.45E-4). We also found significant enrichment for the H3K4me1 histone mark in the germinal matrix (*p* = 6.20E-4), a transient brain region present only during gestational brain development (see **Figure 2** and **Supplementary Table 3** for the magnitude of these enrichments). These annotations represent a subset of eight significantly enriched annotations in ASD, which also included genes conserved in vertebrates, as well as enrichment in histone marks in the anterior caudate and fetal male and female brains. The *cADHD* factor captured 26 significantly enriched annotations which encompassed all of the annotations observed in *uASD* and ASD, albeit with smaller point estimates for the evolutionary annotations. Additional significant annotations for *cADHD* were characterized predominantly by markers for genetic modifications in several brain regions and hormonal centers. A full list of annotations and their relative enrichments is provided in **Supplementary Table 3.**

### T-SEM Uncovers 83 Genes Associated with uASD

We obtained 73 412 gene expression estimates for *uASD* (many of which reflect expression levels for the same gene in different tissues). T-SEM revealed 278 significant hits across 83 unique gene IDs; many of these hits were clustered on chromosomes 8 and 17. These results are visualized as a Miami plot in **Figure 4**. The most highly significant hit corresponded with the downregulation of *PINX1* (z = −5.79, *p* = 6.89E-9), a potent inhibitor of telomerase [36]. The univariate TWAS of ASD revealed 231 significant hits across 69 genes at the same significance threshold used for the *uASD* T-SEM analysis (*p <* 9.37E-5). The *uASD* T-SEM revealed 34 novel gene hits relative to the ASD univariate TWAS. Despite subtracting out shared signal with ADHD, novel genes can arise in this model for genes with particularly discordant effects across ASD and ADHD.

**Figure 3.**
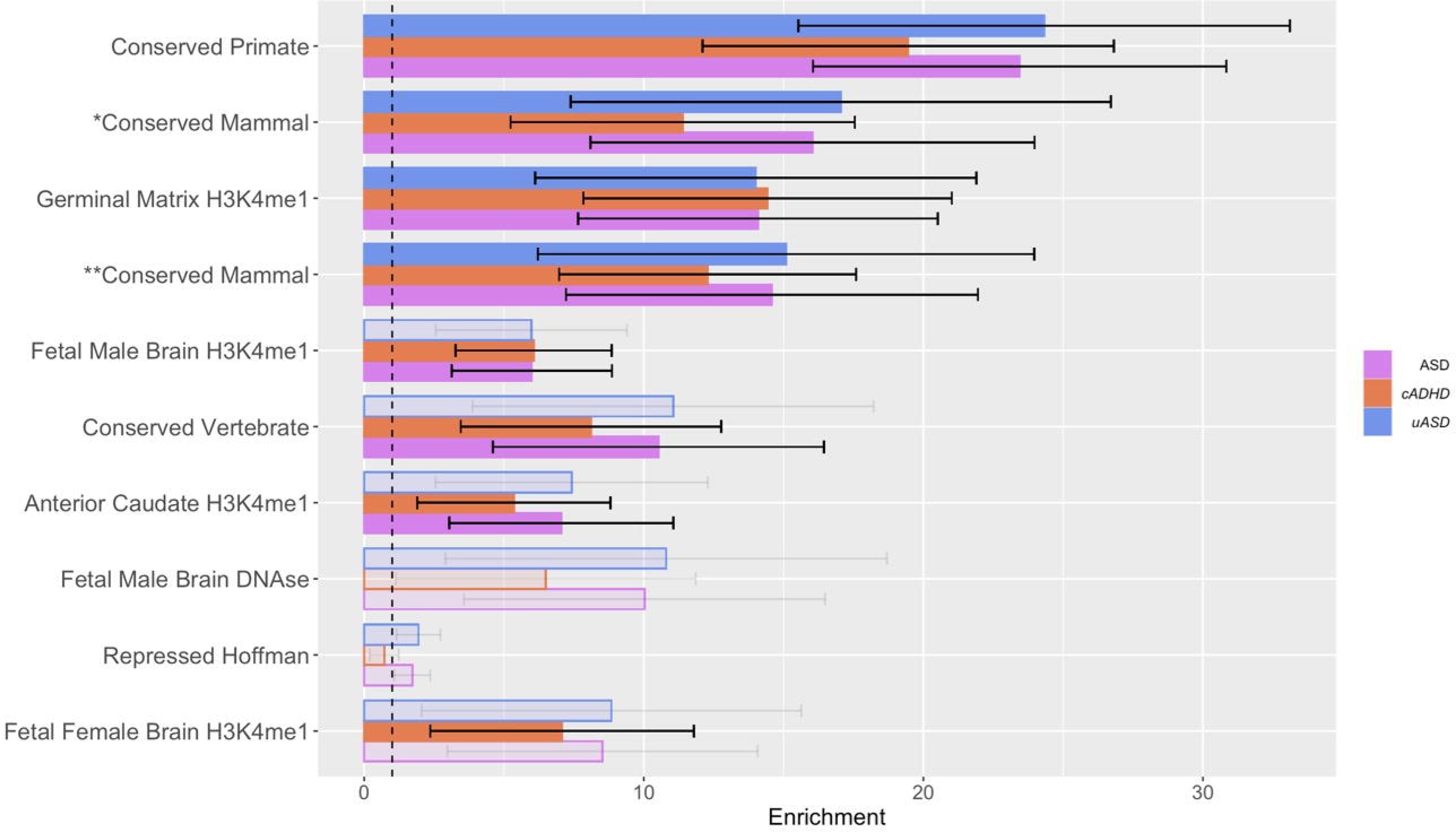
Genetic enrichment of *uASD* for functional annotations. Functional annotations are arranged by ascending *p*-value. Enrichment is measured as the ratio of the proportion of genome-wide relative risk represented by the size of that annotation relative to the entire genome. Null enrichment value is 1.0 (visualized by dashed vertical line), in which the genetic variance captured by that annotation is proportional to the expected genetic variance based on annotation size. Significant enrichments at an FDR threshold are represented by solid blue bars, and error bars represent the 95% CI around the enrichment estimate. For visualization purposes, enrichment values are also provided for ASD (pink) and *cADHD* (red). Single asterisk (*) indicates conserved mammalian genes defined by GERP score. Double asterisk (**) indicates conserved mammalian genes defined by Lindblad-Toh et al. (2011).

**Figure 4.**
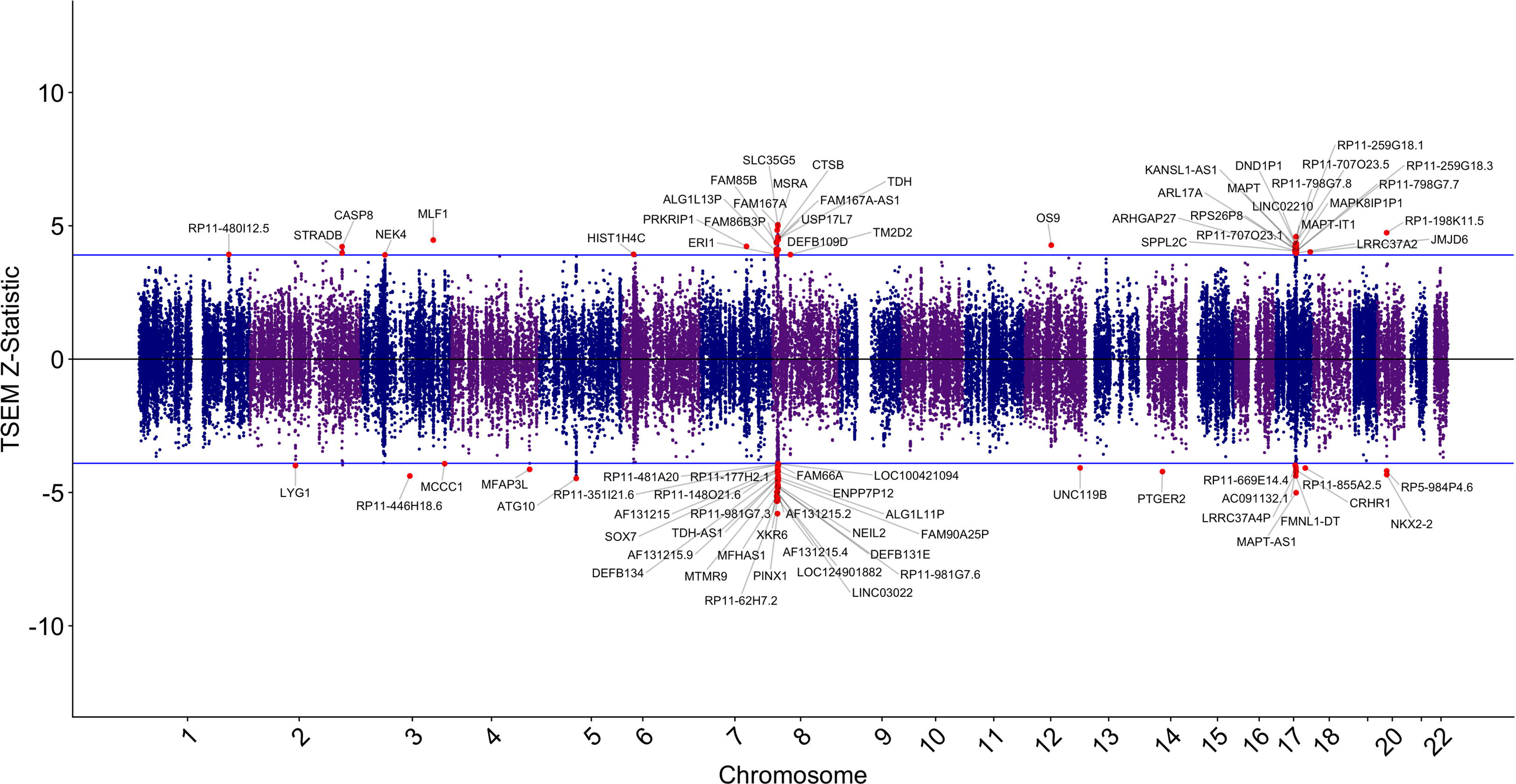
Miami plot of gene expression hits for *uASD*. Upper and lower bounds represent the FDR-adjusted significance threshold (*p* < 9.37E-5). Genes surpassing the upper bound are upregulated and those below the lower bound are downregulated. Significant hits are colored red and labeled with gene ID names.

To identify potential biological pathways implicated in *uASD*, we applied ORA to the significant genes identified by T-SEM. The analysis revealed two gene sets associated with the *uASD* genes; both of which were implicated in skin-related pathologies. The first set relates to bacterial skin diseases (enrichment ratio = 74.53, *p* = 6.55E-9). The second set corresponded to erythema (redness of the skin, often manifesting as a rash) and included all of the genes in the gene list for bacterial skin diseases, with the exception of *FAM167A* (enrichment ratio = 43.27, *p* = 2.20E-6).

## Discussion

The present study leveraged Genomic SEM to dissect the genetic architecture specific to ASD after accounting for shared genetic variance with another neurodevelopmental disorder, ADHD diagnosed in childhood. These ADHD summary statistics evince the highest level of genetic overlap with ASD across the psychiatric space, thereby providing a stringent benchmark for indexing the genetic variance unique to ASD. We find that the majority of genetic variance in ASD, as well as genetic overlap with other clinically relevant traits, is unique from ADHD despite their high levels of genetic overlap [16]. Implementing genome-wide, functional, and gene-expression analyses, we investigated the unique genetic variance at increasing levels of biological granularity. At each level, we interrogated this specific variance and identified distinct biological pathways with specific relevance to ASD.

### Genome-Wide Level

Partitioning the genetic variance unique to ASD, we find a plethora of genetic correlations with cognitive, psychiatric, and other behavioral traits. Notably, we find the strongest correlations between *uASD* and traits related to cognition and education. While the phenotypic literature surrounding ASD and cognitive abilities is mixed [37–39], our findings that *uASD* has a positive genetic association with intelligence and educational attainment corroborate prior genetic studies of ASD [2, 40–42]. Interestingly, we did not see genetic associations with executive function despite meta-analytic research demonstrating ubiquitous executive functioning deficits in ASD [43, 44]. Cognitive traits showed some of the more divergent patterns of association between *uASD* and *cADHD*, with the latter having large and negative genetic associations with the aforementioned cognitive traits. For many cognitive traits, the magnitude of the correlations seen with *uASD* surpassed those seen within ASD broadly. Thus, it appears the genetic variance unique to ASD may have opposing effects to the variance shared between ASD and ADHD, with the unique component driving correlations in the positive direction.

Contrary to cognitive traits, we observed general convergence in the directionality of genetic associations with psychiatric traits, especially in those relating to mood or anxiety disturbances. We therefore conclude that the genetic relationships between ASD and many psychiatric (especially internalizing) phenotypes are not driven solely by the genetic similarity between ASD and ADHD. We also show that ASD is not a simple conglomerate of the genetic components of ADHD and other psychiatric disorders, but rather a genetically distinct construct within the psychiatric space.

### Functional Genomic Level

Functional analyses revealed that the genetic signal unique to ASD was concentrated in evolutionarily conserved genes and the H3K4me1 histone mark in the germinal matrix, a transitory brain region present during the prenatal period which serves as a hub for neural progenitor cells [45]. The enrichment of the H3K4me1 histone mark, indicative of active enhancer elements, reinforces the importance of early epigenetic modifications underlying neurodevelopmental processes in the pathogenesis of ASD [46]. Moreover, our observation of this unique enrichment underscores the value of employing multivariate genomic analyses to dissect disorder-specific biological pathways, facilitating a deeper understanding of the molecular mechanisms underlying ASD etiology.

### Transcriptome-Wide Level

Transcriptome-wide analyses revealed 83 unique genes with differential expression linked with ASD independent of ADHD. Notably, we find several novel gene hits unique to ASD, reflecting genes with highly discordant effects across ASD and ADHD. Focusing on genes with expression associated with *uASD*, we find overlap with gene sets implicated in two classes of skin-related pathologies: bacterial skin disease and erythema (non-specific reddening of the skin). There are well-documented links between ASD and various immune-mediated conditions, with medical research focusing on the association with atopic dermatitis (i.e., eczema) [47–50]. Dysfunction of the immune system and inflammatory processes have been hypothesized to contribute jointly to ASD and skin disorders such as atopic dermatitis [51]; however, there is a dearth of literature apart from the current study which has explicitly identified gene clusters implicated in both syndromes.

### Limitations

The current analyses were restricted to GWAS summary statistics derived exclusively from individuals of European ancestry [52], which hinders the generalizability of our findings due to differences in allele frequency and LD structure across ancestrally diverse populations [53]. Efforts to broaden representation in GWAS analyses are crucial for extending genetic insights to other ancestral groups [54]. It is also critical to recognize that ASD is a highly heterogeneous condition characterized by a broad range of clinical profiles and severity levels [55, 56]. While these diverse phenotypes of ASD are likely attributable to different genetic backgrounds [2], the use of a general ASD GWAS restricts our ability to parse any phenotypic heterogeneity in the sample. Furthermore, many behavioral and psychiatric traits, including ASD, show discrepancies between family- and SNP-based estimates of heritability [57], with a component of ASD’s missing heritability thought to be driven by rare, high-impact variants undetected by GWASs [58–60]. It is therefore important to recognize that we are only examining a subset of the genetic factors thought to independently contribute to ASD etiology.

### Conclusions

Taken together, we provide insights across multiple levels of biology that characterize the genetic signal unique to ASD. Relative to ADHD, we find evidence for divergent patterns of relationships with a range of clinically relevant correlates (e.g., cognition) along with unique patterns of functional enrichment and gene expression that implicate neurobiological processes and disease states linked to ASD in the extant literature. While ASD has often been discussed as unique within the psychiatric space, the current findings clarify and characterize the biological substrata that differentiate this complex neuropsychiatric disorder.

## Data Availability

All data used in these analyses is publicly available. GWAS summary statistics for autism spectrum disorder can be downloaded at https://figshare.com/articles/dataset/asd2019/14671989. GWAS summary statistics for childhood-diagnosed attention-deficit/hyperactivity disorder can be downloaded at https://ipsych.dk/en/research/downloads/.

## Acknowledgments

We are extremely grateful for the efforts put forth by the investigators and participants from data resources such as the Psychiatric Genetics Consortium, iPSYCH, and UK Biobank for making these analyses possible.

