## Supplementary Figure for "Characterizing Genetic Pathways Unique to Autism Spectrum Disorder at Multiple Levels of Biological Analysis"

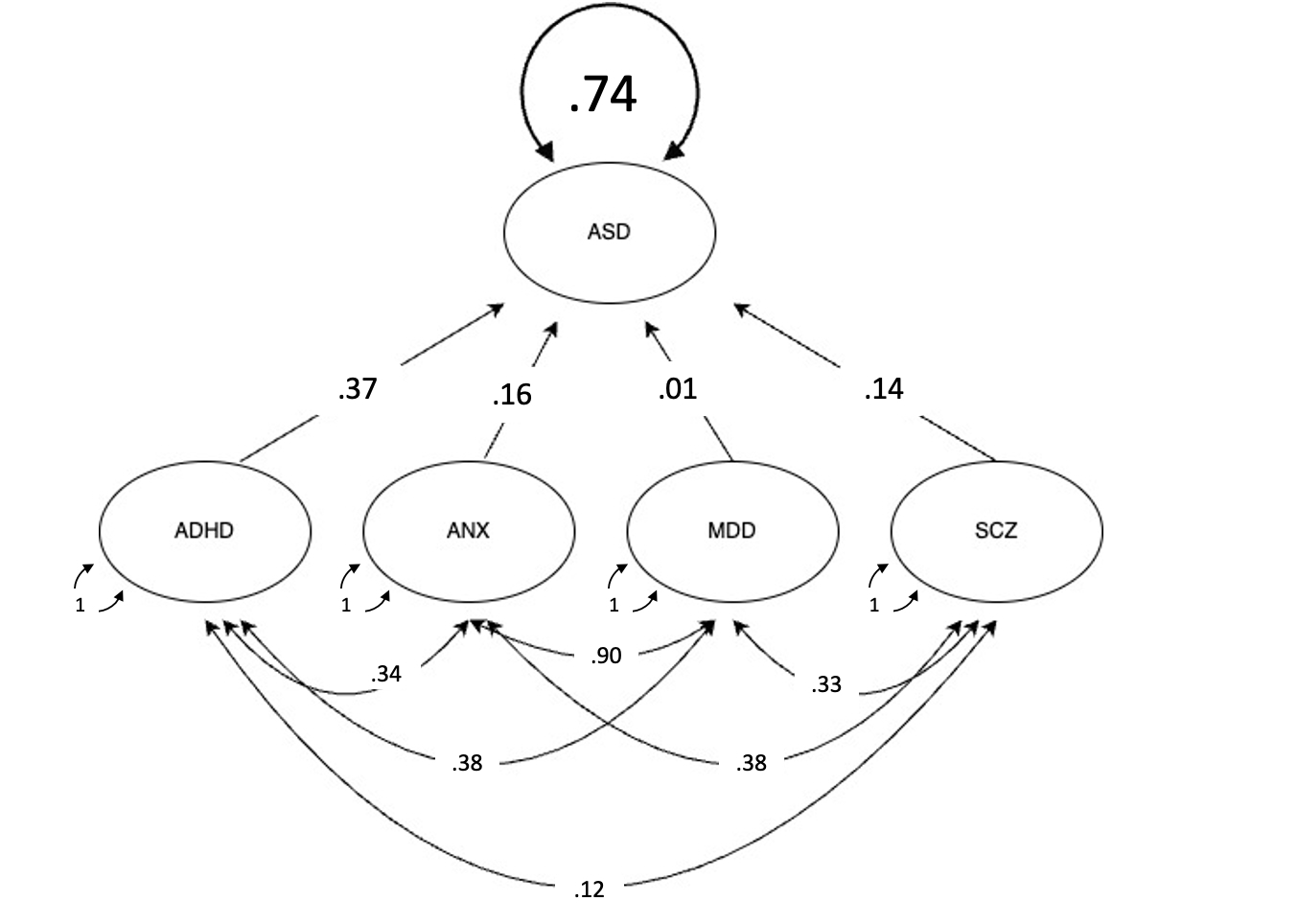

**Supplementary Figure 1.** **Note.** Path diagram showing standardized path estimates for multiple regression model estimating the residual genetic variance in ASD after accounting for shared genetic variance with four correlated psychiatric disorders as predictors: childhood-diagnosed ADHD, anxiety (ANX), major depressive disorder (MDD), and schizophrenia (SCZ).
